# Investigation of an ongoing outbreak of ceftriaxone-resistant *Salmonella enterica* serovar Typhi in Bangladesh

**DOI:** 10.1101/2024.12.24.24319600

**Authors:** Yogesh Hooda, Arif Mohammad Tanmoy, Sudipta Deb Nath, Anannya Barman Jui, Al Amin, Hafizur Rahman, Neoyman Nasir Shorkar, Naito Kanon, Md Asadur Rahman, Denise O Garrett, Mohammad Shahidul Islam, ASM Nawshad Uddin Ahmed, Samir K Saha, Senjuti Saha

## Abstract

We report an outbreak of ceftriaxone-resistant *Salmonella* Typhi in Bangladesh, with 47 cases identified from April-September 2024. Isolates belong to genotype 4.3.1.2 and harbor the *bla*_CTX- M-15_ gene on the pCROB1 plasmid. This unique genotype-plasmid lineage represents a recent introduction, necessitating enhanced surveillance, antimicrobial stewardship, and consideration of vaccination strategies.

## Main text

*Salmonella* Typhi, the causative agent of typhoid fever, remains a major public health concern, particularly in South Asia, which accounts for approximately 70% of global cases [1]. Reports of drug-resistant *Salmonella* Typhi have increased in recent decades. Of particular concern are strains resistant to ceftriaxone and azithromycin [2]. In 2016, an outbreak of extensively drug-resistant (XDR) *Salmonella* Typhi was identified in Pakistan [3]. These strains are resistant to chloramphenicol, ampicillin, cotrimoxazole, fluoroquinolones, and third generation cephalosporins [3]. Sporadic cases of independent acquisition of ceftriaxone resistance have also been previously reported from Bangladesh [4], India [5], and UK [6]. The high use of ceftriaxone for empirical treatment in South Asia underscores the need for public health authorities to remain vigilant against the emergence and spread of ceftriaxone-resistant *Salmonella* Typhi [2].

In Bangladesh, *Salmonella* Typhi is the most common cause of bloodstream infections in children older than two months [7]. However, most previous surveillance studies have primarily focused on Dhaka, the capital city [7]. To address this gap and monitor the burden of disease and associated antibiotic susceptibility patterns nationwide, we began expanding our surveillance network in January 2023 to include clinics affiliated with the Popular Diagnostic Centre Ltd (PDCL) (Figure 1A). As of March 2024, this network encompasses 20 clinics across 11 districts, including 10 in Dhaka (Figure 1A). Leveraging this expanded passive surveillance for typhoid fever, we report data from January 2023 through September 2024, describing the emergence, spread, and genomic epidemiology of a ceftriaxone-resistant clone of *Salmonella* Typhi in Bangladesh. Ethical approval for this study was obtained from the Ethics Review Boards of the Child Health Research Foundation (CHRF) and the Bangladesh Shishu Hospital and Institute.

**Figure 1:**
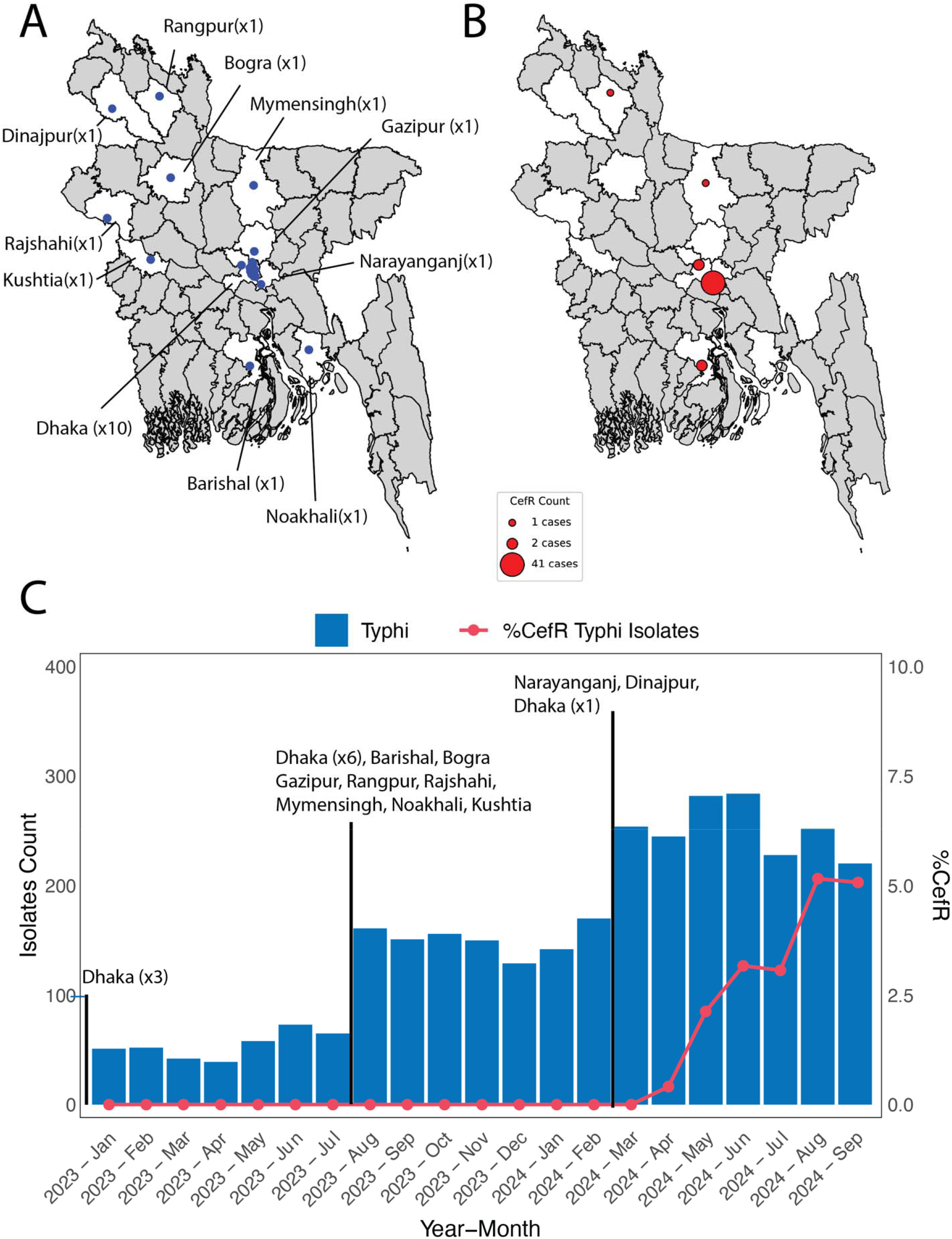
Outbreak of ceftriaxone-resistant *Salmonella* Typhi in Bangladesh. A) Locations of the 20 clinics across 11 districts in white included in this study. Each clinic location is shown as a blue dot. Districts not included in this study are colored in grey. B) Geographical distribution of ceftriaxone resistant (CefR) *Salmonella* Typhi isolates across Bangladesh. The size of each bubble represents the number of isolates per district. C) Timeline of the surveillance network, showing the inclusion of clinics and the number of *Salmonella* Typhi isolates detected. The rate of CefR *Salmonella* Typhi among all *Salmonella* Typhi cases is highlighted in red. This surveillance study began in January 2023 with three clinics in Dhaka. In August 2023, six additional clinics in Dhaka were added, along with clinics in Barisal, Bogra, Gazipur, Kushtia, Rangpur, Rajshahi, Mymensingh, and Noakhali. By March 2024, one more clinic in Dhaka, along with clinics in Narayanganj and Dinajpur, were incorporated.

### The Study

PDCL clinics are outpatient facilities that perform blood cultures for patients as advised by their treating physicians. If a blood culture yields *Salmonella* Typhi, the isolate is transported to the CHRF laboratory for serovar confirmation and antibiotic susceptibility testing. These processes follow established methodologies, including biochemical and slide-agglutination tests (*Salmonella* agglutinating antisera; Thermo Scientific, MA, USA) and CLSI-guided Kirby-Bauer disc diffusion methods (Oxoid, Thermo Scientific, MA, USA) [8].

The first ceftriaxone resistant (CefR) *Salmonella* Typhi isolate was identified on April 27, 2024, at the PDCL clinic in Narayanganj, a city located 30 km from Dhaka. Subsequent monthly detection of CefR strains continued through the study period, culminating in a total of 47 cases by September 2024. Of these, 41 cases were from Narayanganj, with the remaining six from other districts: Dhaka (n = 2), Barisal (n = 2), Mymensingh (n = 1), and Rangpur (n = 1) (Figure 1B). The isolation rate of CefR *Salmonella* Typhi increased from 0% in March 2024 to over 5% of all isolates in September 2024 (Figure 1C).

All 47 isolates demonstrated resistance to amoxicillin and ceftriaxone, non-susceptibility to fluoroquinolones, but sensitivity to chloramphenicol, cotrimoxazole, azithromycin, and meropenem (Table 1).

**Table 1.**
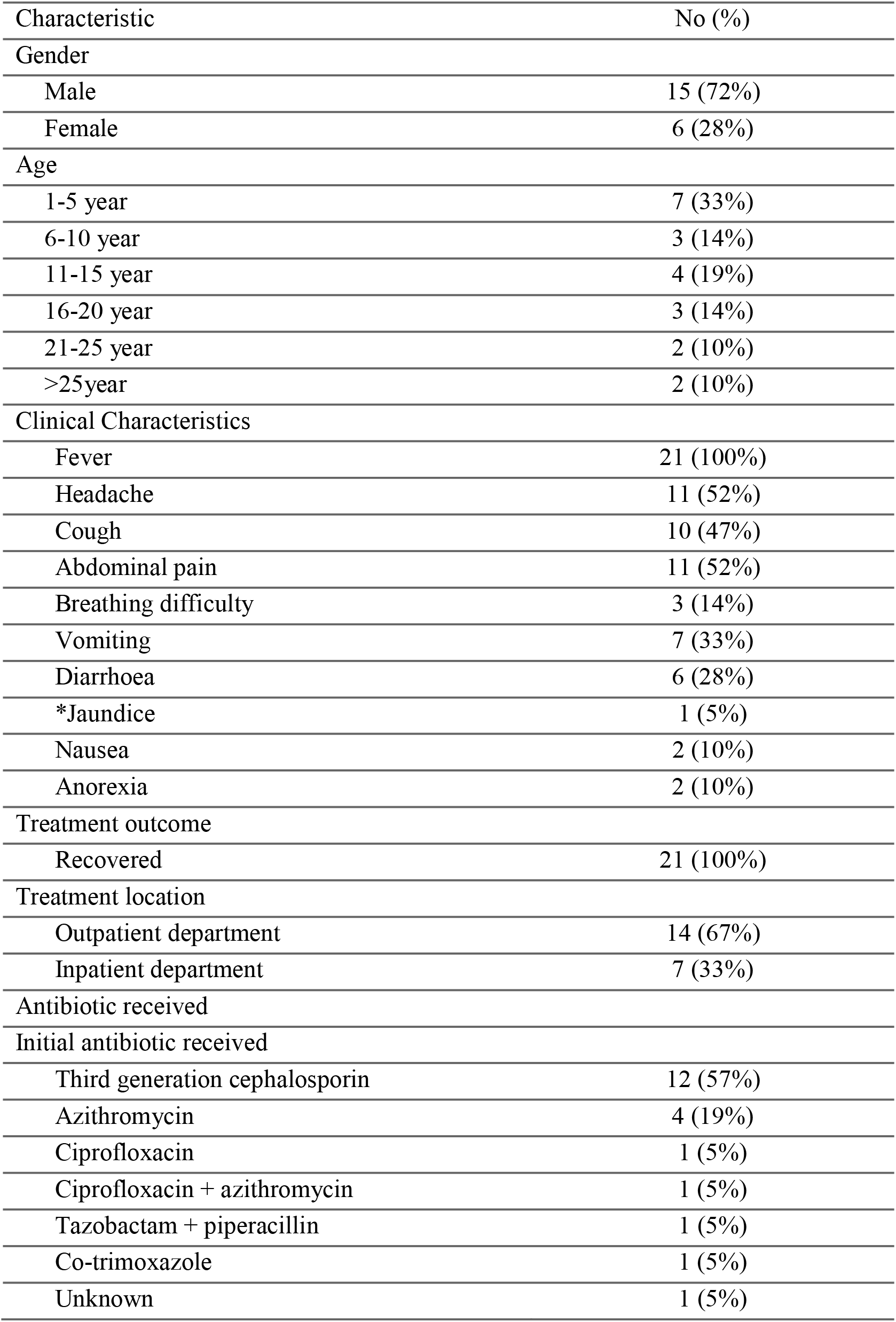
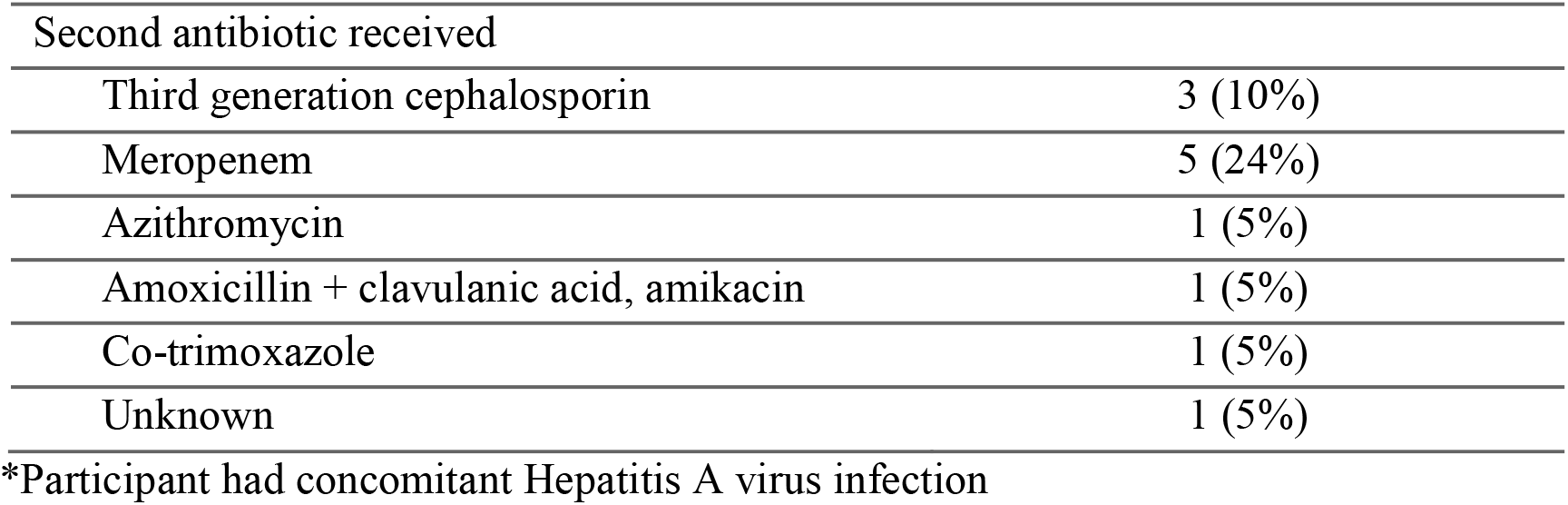
Clinical, demographic, and epidemiological features of patients (n = 21) with ceftriaxone resistant *Salmonella* Typhi infections.

To investigate the basis of ceftriaxone resistance, 17 of the 47 strains were subjected to whole genome sequencing. These included 14 isolates from Narayanganj and one each from Barishal, Mymensingh, and Rangpur. Libraries were prepared using the NEB library preparation kit (NEB, Ipswich, MA, USA) and sequenced on a NextSeq2000 platform (2×150 bp). Raw fastq files have been submitted to European Nucleotide Archive under accession, ERP167392. Genome assembly was performed using Unicycler v0.5.1 [9] and the resulting contigs were analyzed with Mykrobe [10]. All 17 CefR genomes were classified as genotype 4.3.1.2. Ceftriaxone resistance was conferred by the *bla*_CTX-M-15_ gene, which exhibited 100% sequence identity to the gene found in XDR *Salmonella* Typhi strains from Pakistan (genotype 4.3.1.1.P1) [3]. Phylogenetic analysis was conducted using Bowtie2, Samtools, Gubbins, and RAxML, following the pipeline described by Tanmoy *et al* [11].

An IncY plasmid was identified in all 17 sequenced strains using Mykrobe, and further characterized using plasmidSPAdes v3.15.5 [14]. One of the isolates, STY_0313, produced a plasmid sequence measuring 103.9 kbp. This IncY plasmid, referred to as “pCROB1,” carried the *bla*_CTX-M-15_ gene and appeared to encode a phage element (Figure 2). A similar phage-plasmid associated with genotype 4.3.1.1 was previously identified by Public Health England among travel-related typhoid cases from Iraq [6]. These findings suggest that a unique genotype- plasmid lineage underpins the ongoing ceftriaxone resistance outbreak in Bangladesh.

**Figure 2:**
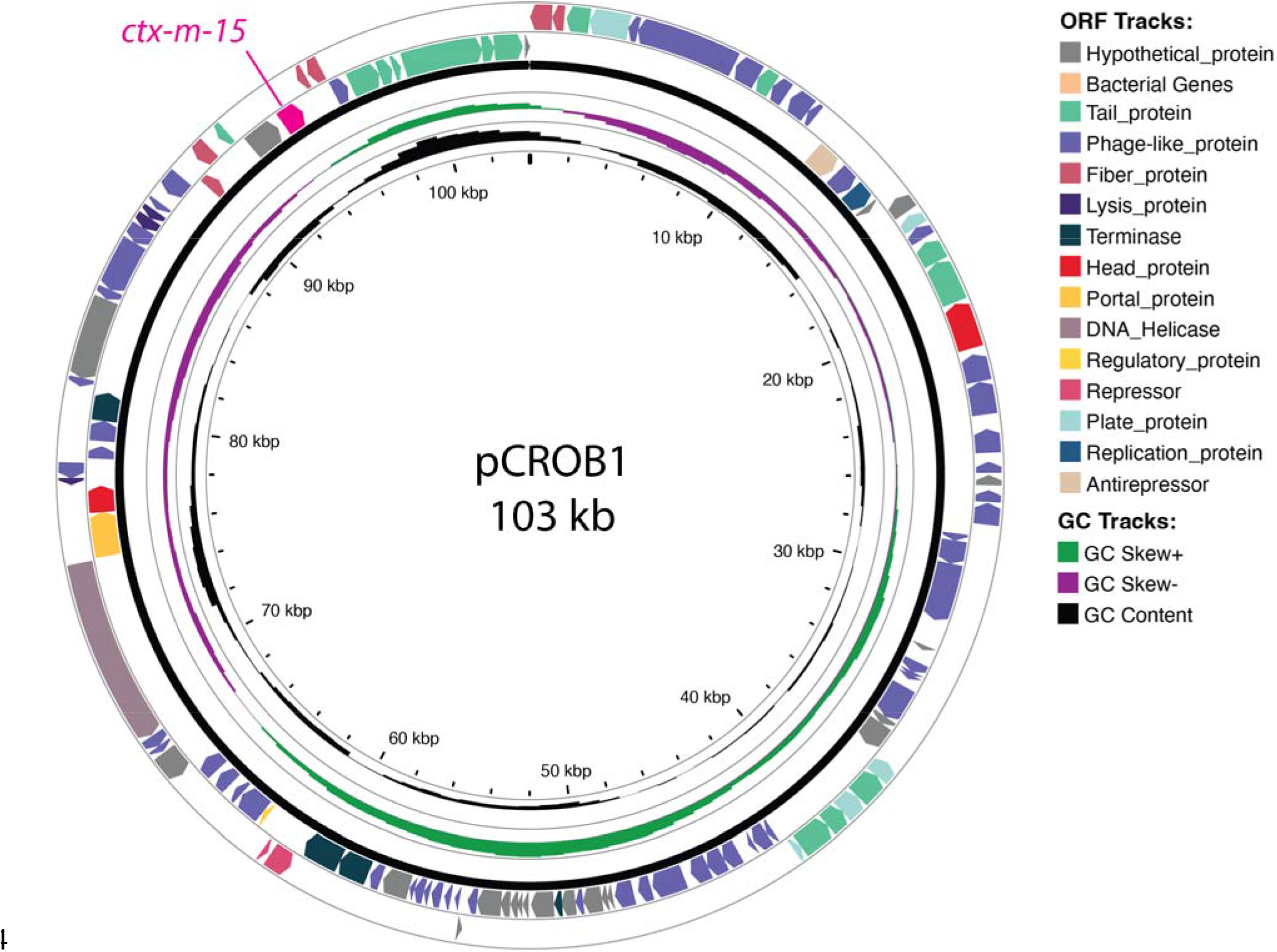
Annotated plasmid map of pCROB1. This plasmid is closely related to one previously described by Public Health England [6]. It harbors the *bla*_CTX-M-15_ gene and includes an intact phage element, as identified using PHASTEST [12]. Tracks around the plasmid map highlight GC-content and predicted open reading frames (ORFs).

Genotype 4.3.1.2 is rarely identified in Bangladesh, accounting for only 0.5% (7/1,356) of all *Salmonella* Typhi whole-genome sequences from 1999 to 2018 [15] but it is more commonly found in India and Nepal (Figure 3). To investigate its phylogenetic context, we constructed a tree using global database sequences from genotype 4.3.1 and its subtypes, with a focus on 4.3.1.2 and its sub-lineages. The recent ceftriaxone-resistant isolates from Bangladesh are closely related to strains from India and Nepal, rather than earlier 4.3.1.2 isolates from Bangladesh (Figure 3). However, none of these strains from India or Nepal were reported to be ceftriaxone-resistant or carry the pCROB1 plasmid.

**Figure 3:**
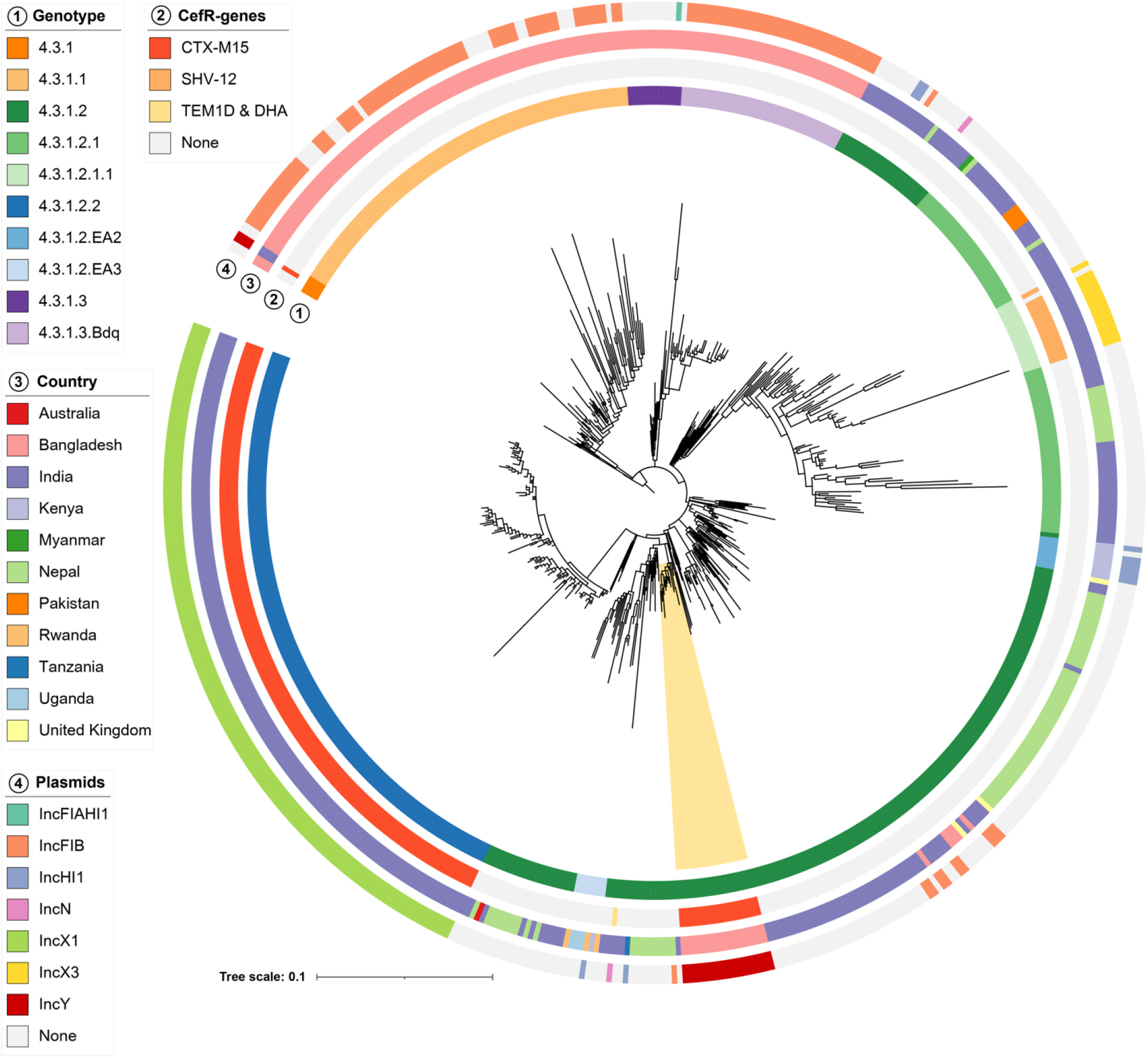
Phylogenetic tree of *Salmonella* Typhi genotype 4.3.1, including 17 recent ceftriaxone-resistant strains from Bangladesh. The CefR strains sequenced in this study are highlighted in yellow. The tree displays different ceftriaxone resistance genes, countries of isolation, and associated plasmid elements. For context, 529 genomes from genotype 4.3.1 and its subtypes were selected from Pathogenwatch and other studies conducted in Bangladesh, India, and Pakistan [13].

We propose naming this distinct subclade “lineage 4.3.1.2.B1”. While ceftriaxone resistance has been reported in genotype 4.3.1.2 strains from India [5], the molecular basis of resistance in lineage 4.3.1.2.B1 differs, suggesting an independent acquisition of the ceftriaxone-encoding phage-plasmid by this genotype.

To better understand the disease history, treatment, outcomes, and travel history of patients with ceftriaxone-resistant typhoid, 35 cases identified till August 31, 2024, were selected for telephone interviews, with 21 (60%) successfully completed. Of these, 15 (72%) were male, and 15 (72%) were under 18 years of age. All patients presented with fever; additional symptoms included headaches (52%), abdominal pain (52%), and cough (47%). Fourteen (66%) were treated as outpatients, while seven (33%) required hospitalization. Most patients (13/21; 65%) initially received second- or third generation cephalosporins, with 45% (6/13) switching to azithromycin (n = 1), cotrimoxazole (n = 1), or meropenem (n = 3) based on susceptibility results. One patient could not recall the second antibiotic used. All patients recovered, with an average illness duration of 19 days (range: 12–28 days). None of the interviewed patients reported travel outside Bangladesh, suggesting local circulation of this strain. Three cases occurred outside Narayanganj, the initial outbreak site, and two had no travel history to Narayanganj within 15 days of illness onset, indicating potential spread to other districts.

## Discussion

This outbreak highlights the urgent need for robust surveillance systems to detect and respond to the emergence of drug-resistant *Salmonella* Typhi. The unique lineage 4.3.1.2.B1 carrying the *bla*_CTX-M-15_ gene on the pCROB1 plasmid represents a concerning development, given the potential for regional/international dissemination.

Given the potential for azithromycin resistance to emerge in lineage 4.3.1.2.B1, healthcare systems may need to prepare for a shift back to older antibiotics, such as cotrimoxazole and chloramphenicol. Notably, 4.3.1.2.B1 strains remain sensitive to cotrimoxazole and chloramphenicol, indicating these first-line antibiotics may serve as viable alternatives to azithromycin and meropenem. One patient successfully recovered with cotrimoxazole, emphasizing its continued relevance in treatment.

Our findings also underscore the role of widespread ceftriaxone use in selecting for resistant strains. Public health efforts should focus on antimicrobial stewardship, and public education on appropriate antibiotic use. In addition, efforts should be strengthened towards implementing and promoting immunization with typhoid conjugate vaccines (TCVs) to curb further spread of resistant strains.

## Conclusion

We describe a significant outbreak of ceftriaxone-resistant *Salmonella* Typhi in Bangladesh, driven by a unique genotype-plasmid lineage. As ceftriaxone remains a cornerstone of empirical treatment for typhoid, its declining efficacy is of great public health concern.

Public health authorities should strengthen surveillance and prioritize vaccination to prevent further dissemination. Additionally, global health systems should remain vigilant for travel- related cases. Ongoing genomic investigations are essential to track resistance mechanisms and inform policy decisions. Proactive measures are imperative to mitigate the public health threat posed by emerging AMR pathogens.

## Data Availability

All data in the present study are contained in the manuscript.

## Notes

### Competing Interest Statement

The authors have declared no competing interest.

### Funding Statement

The study was funded by the Bill and Melinda Gates Foundation grant to SS (INV-051975).

### Author Declarations

Ethical approval for this study was obtained from the Ethics Review Boards of the Child Health Research Foundation (CHRF) and the Bangladesh Shishu Hospital and Institute.

## References

1. GBD 2017 Typhoid and Paratyphoid Collaborators. The global burden of typhoid and paratyphoid fevers: a systematic analysis for the Global Burden of Disease Study 2017. Lancet Infect Dis 2019;

2. Hooda Y, Tanmoy AM, Sajib MSI, Saha S. Mass azithromycin administration: considerations in an increasingly resistant world. BMJ Global Health 2020; 5:e002446.

3. Klemm EJ, Shakoor S, Page AJ, et al. Emergence of an Extensively Drug-Resistant Salmonella enterica Serovar Typhi Clone Harboring a Promiscuous Plasmid Encoding Resistance to Fluoroquinolones and Third-Generation Cephalosporins. mBio 2018; 9:e00105–18.

4. Djeghout B, Saha S, Sajib MSI, et al. Ceftriaxone-resistant Salmonella Typhi carries an IncI1-ST31 plasmid encoding CTX-M-15. Journal of Medical Microbiology 2018; 67:620–627.

5. Argimón S, Nagaraj G, Shamanna V, et al. Circulation of Third-Generation Cephalosporin Resistant Salmonella Typhi in Mumbai, India. Clinical Infectious Diseases 2022; 74:2234–2237.

6. Godbole G, McCann N, Jones SM, Dallman TJ, Brown M. Ceftriaxone-resistant Salmonella Typhi in a traveller returning from a mass gathering in Iraq. The Lancet Infectious Diseases 2019; 19:467.

7. Saha S, Islam M, Uddin MJ, et al. Integration of enteric fever surveillance into the WHO-coordinated Invasive Bacterial-Vaccine Preventable Diseases (IB-VPD) platform: A low cost approach to track an increasingly important disease. PLOS Neglected Tropical Diseases 2017; 11:e0005999.

8. Tanmoy AM, Hooda Y, Sajib MSI, et al. Trends in antimicrobial resistance amongst Salmonella Typhi in Bangladesh: A 24-year retrospective observational study (1999–2022). PLOS Neglected Tropical Diseases 2024; 18:e0012558.

9. Wick RR, Judd LM, Gorrie CL, Holt KE. Unicycler: Resolving bacterial genome assemblies from short and long sequencing reads. PLOS Computational Biology 2017; 13:e1005595.

10. Ingle DJ, Hawkey J, Hunt M, et al. Typhi Mykrobe: fast and accurate lineage identification and antimicrobial resistance genotyping directly from sequence reads for the typhoid fever agent Salmonella Typhi. 2024; :2024.09.30.613582. Available at: https://www.biorxiv.org/content/10.1101/2024.09.30.613582v1. Accessed 3 November 2024.

11. Tanmoy AM, Hooda Y, Sajib MSI, et al. Paratype: a genotyping tool for Salmonella Paratyphi A reveals its global genomic diversity. Nat Commun 2022; 13:7912.

12. Wishart DS, Han S, Saha S, et al. PHASTEST: faster than PHASTER, better than PHAST. Nucleic Acids Research 2023; 51:W443–W450.

13. Argimón S, David S, Underwood A, et al. Rapid Genomic Characterization and Global Surveillance of Klebsiella Using Pathogenwatch. Clinical Infectious Diseases 2021; 73:S325–S335.

14. Antipov D, Hartwick N, Shen M, Raiko M, Lapidus A, Pevzner PA. plasmidSPAdes: assembling plasmids from whole genome sequencing data. Bioinformatics 2016; 32:3380–3387.

15. Carey ME, Dyson ZA, Ingle DJ, et al. Global diversity and antimicrobial resistance of typhoid fever pathogens: Insights from a meta-analysis of 13,000 Salmonella Typhi genomes. eLife 2023; 12:e85867.

